# General Model for COVID-19 Spreading with Consideration of Intercity Migration, Insufficient Testing and Active Intervention: Application to Study of Pandemic Progression in Japan and USA

**DOI:** 10.1101/2020.03.25.20043380

**Authors:** Choujun Zhan, Chi K. Tse, Zhikang Lai, Xiaoyun Chen, Mingshen Mo

## Abstract

A new Susceptible-Exposed-Infected-Confirmed-Removed (SEICR) model with consideration of intercity travel and active intervention is proposed for predicting the spreading progression of the 2019 New Coronavirus Disease (COVID-19). The model takes into account the known or reported number of infected cases being fewer than the actual number of infected individuals due to insufficient testing. The model integrates intercity travel data to track the movement of exposed and infected individuals among cities, and allows different levels of active intervention to be considered so that realistic prediction of the number of infected individuals can be performed. The data of the COVID-19 infection cases and the intercity travel data for Japan (January 15 to March 20, 2020) and the USA (February 20 to March 20, 2020) are used to illustrate the prediction of the pandemic progression in 47 regions of Japan and 50 states (plus a federal district) in the USA. By fitting the model with the data, we reveal that, as of March 19, 2020, the number of infected individuals in Japan and the USA could be twenty-fold and five-fold as many as the number of confirmed cases, respectively. Moreover, the model generates future progression profiles for different levels of intervention by setting the parameters relative to the values found from the data fitting. Results show that without tightening the implementation of active intervention, Japan and the USA will see about 6.55% and 18.2% of the population eventually infected, and with drastic ten-fold elevated active intervention, the number of people eventually infected can be reduced by up to 95% in Japan and 70% in the USA. Finally, an assessment of the relative effectiveness of active intervention and personal protective measures is discussed. With a highly vigilant public maintaining personal hygiene and exercising strict protective measures, the percentage of population infected can be further reduced to 0.23% in Japan and 2.7% in the USA.

## 1. Introduction

The global spread of the 2019 New Coronavirus Disease (COVID-19) has shown no sign of subsiding since its emergence in Wuhan, China, in December 2019 [1]. As of March 21, 2020, a total of 276,472 cases of COVID-19 infection have been confirmed in over 185 countries, with a death toll of 11,417 [2]. Different control strategies at different levels of stringency have been applied to slow the spread of the virus in different countries [3]. While some countries have seen peaks of infected cases and observed significant reduction in the number of new infections in the local communities [2, 4], the spreading has continued in many countries, and surges in infected cases have been observed in Europe, USA and Australia. Intercity travel has been found to be a contributing factor to the rapid spread of the virus [5, 6]. Thus, effective models for describing the pandemic progression in different cities should take into consideration the volume of intercity travels. Furthermore, the rapid spread of the virus in a population has often been a result of delayed information or unawareness of the real situation in that population, despite the wide dissemination of information related to COVID-19 outbreaks in other parts of the world. The most notable information latency lies in the number of confirmed cases reported, which depends on the ability of the particular country or city to perform tests as well as the possible bureaucracy in the local system of reporting. Thus, the number of confirmed cases is almost certainly not the true number of infected individuals at any given time [7], and an improved model for predicting the spreading progression should incorporate the latency associated with the reporting system as well as the possible missing cases leading to delay and loss of information. The traditional Susceptible-Exposed-Infectious-Recovered (SEIR) model [8, 9] thus has obvious shortfalls in describing the spreading dynamics of COVID-19 pandemic. In this work, we attempt to fill the main gap between the number of confirmed cases and the actual number of infected cases. Specifically, in the proposed model, an infected individual may become a confirmed case and then recovered/removed. Moreover, an infected individual may also be recovered/removed without being confirmed as infected. In other words, the basic model proposed here is a Susceptible-Exposed-Infectious-**Confirmed**-Recovered (SEICR) model, which has an additional state corresponding to an individual having been confirmed by the authority as being infected.

On the basis of an SEICR model, we develop a model incorporating intercity travel data which accounts for any increase or decrease in the number of exposed and infected individuals in a city due to intercity migration. Furthermore, the level of intervention in the form of travel restriction, regional lockdown or other active control measures would profoundly influence the rapidity of the spread of the virus and the eventual number of infected cases. The model should therefore allow the level of active intervention to be included as a control parameter and produce the appropriate progression profile. A specific parameter is used to adjust the level of active intervention in the simulation of future progression profiles, which corresponds quantitatively to the increase in the number of individuals eventually infected due to an additional infected individual at any given time.

In this work, we apply the model to study the COVID-19 spreading progression in Japan and the USA. Data of confirmed and recovered cases in 47 Japanese prefectures or regions (January 15 to March 20, 2020) and 51 states in the USA including Washington DC (February 20 to March 20, 2020) are used for fitting with the model and retrieval of parameter values. The parameters found are then adjusted to produce future progression trajectories corresponding to the implementation of different levels of active intervention. From the set of best-fit parameters, we reveal that the actual number of infected individuals could be up to 20-fold and 5-fold as many as the confirmed numbers in Japan and the USA, respectively, as of March 19, 2020. Furthermore, if the level of active intervention is kept unchanged, the percentage of population eventually infected in Osaka-fu and Tokyo-to will reach around 12% and 4.2% of the population (around 2,300,000 and 600,000 people), respectively, and in total, Japan will have about 6.55% of its population eventually infected. However, implementing a four-fold elevated active intervention will improve the situation substantially, with the percentage of population infected in Osaka-fu and Tokyo-to reduced to 4.2% and 2.3%, respectively, and over 75% reduction in the overall number of infected cases in Japan. Our results for the USA also show a rather detrimental situation if the level of active intervention remains at the status quo in the coming months, with California and New York state having around 15% and 37.5% of their population (5,800,000 and 7,300,000 people) eventually infected, respectively, and the overall infected population will reach 18.2%. However, by implementing four-fold elevated active intervention, the percentage of infected population in the USA can be reduced to 9.3%. Furthermore, the effectiveness of the public in exercising protective measures can be assessed by varying the infection rates in the model, and it is found that active government intervention is more effective for Japan, while exercising protective measures by the public is more important for the USA. With the public raising its level of vigilance in exercising strict protective measures and the government drastically elevating its active intervention, the percentage of population getting infected can be reduced to 0.23% in Japan and 2.7% in the USA.

## 2. Data

The World Health Organization currently sets the alert level of COVID-19 to the highest, and has made data related to the pandemic available to the public in a series of situation reports as well as other formats [10]. Our data include the number of confirmed infected cases, the cumulative number of confirmed infected cases, the number of recovered cases, and death tolls, for 47 individual prefectures and regions in Japan, from January 15 to March 20, 2020, and for 50 states and a federal district (Washington DC) in the USA, from February 20 to March 20, 2020. Data organized in convenient formats are also available elsewhere [2, 11, 12]. Moreover, the monthly intercity migration data for February 2020 are available from official statistics provided by the Japanese government [13], and are used as indicative migration strengths between prefectures or regions in Japan. For the USA, annual data for the volume of inter-state travellers are available from the Census Bureau [14] and the Bureau of Transportation Statistics [15].

## 3. Method

### 3.1. Migration-Data Augmented SEICR Model

In the proposed Susceptible-Exposed-Infectious-Confirmed-Recovered (SEICR) model, each individual would assume one of five possible states at any time, namely, susceptible (*S*), exposed (*E*), infected (*I*), confirmed (*C*), and recovered/removed (*R*). Compared to the traditional SEIR model [8, 9], the new SEICR model has an additional *C* state, corresponding an individual having been confirmed by the authority as infected. Thus, not all infected individuals will become confirmed, and some infected individuals will transit to the recovered state without going through the confirmed state. For city or region *j*, the number of individuals in the five states are *S*_*j*_(*t*), *E*_*j*_(*t*), *I*_*j*_(*t*), *C*_*j*_(*t*) and *R*_*j*_(*t*), at time *t*. The transitions of the five states are illustrated in Figure 1. In addition, *P*_*j*_(*t*) stands for the population of region *j*. Furthermore, to account for intercity movement, we introduce a migration strength, *m*_*ij*_(*t*), which represents an indicative volume of people moving from region *i* to region *j* at time *t* [4]. Then, the augmented SEICR model is given as follows:

**Figure 1:**
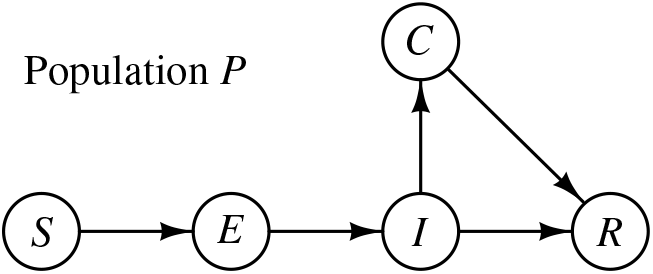
State transition flow chart.

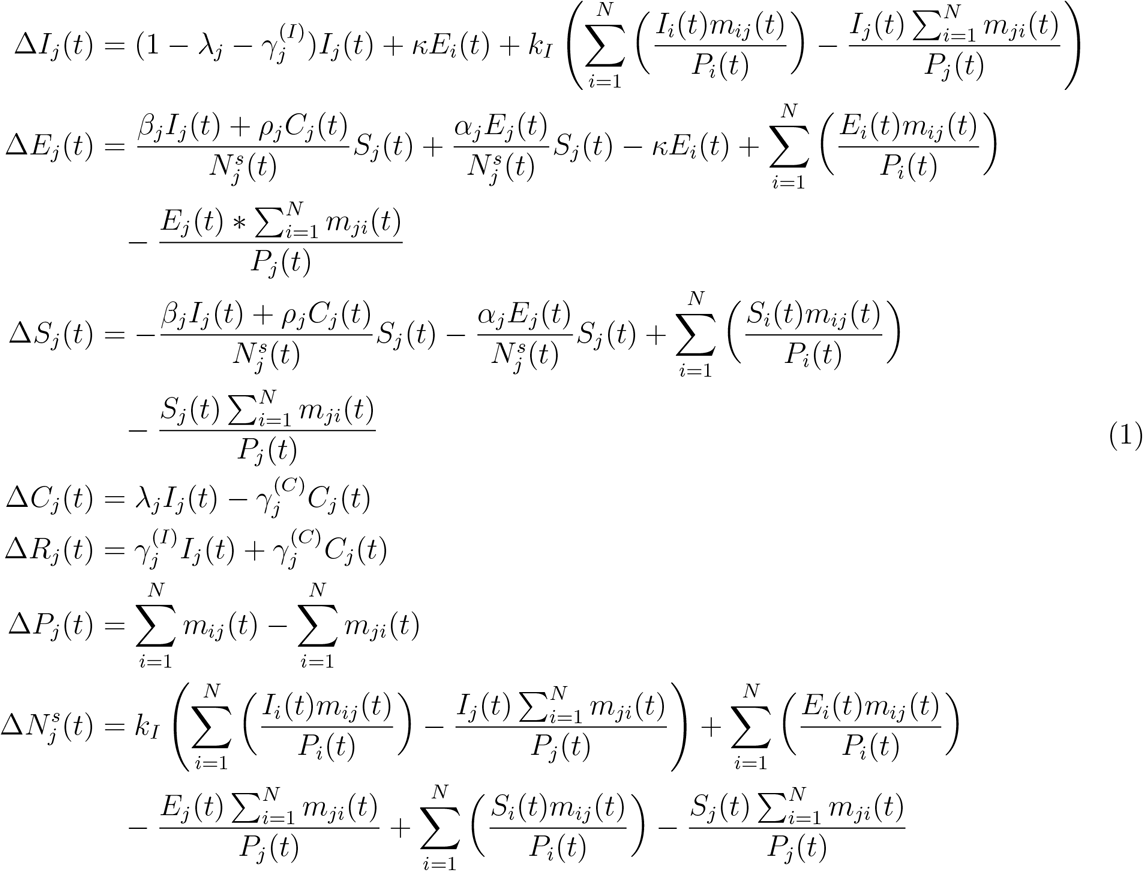

where Δ*I*_*j*_(*t*) = *I*_*j*_(*t* + 1) *− I*_*j*_(*t*), Δ*E*_*j*_(*t*) = *E*_*j*_(*t* + 1) *− E*_*j*_(*t*), Δ*S*_*j*_(*t*) = *S*_*j*_(*t* + 1) *− S*_*j*_(*t*), Δ*C*_*j*_(*t*) = *C*_*j*_(*t* + 1) *− C*_*j*_(*t*), Δ*R*_*j*_(*t*) = *R*_*j*_(*t* + 1) *− R*_*j*_(*t*), 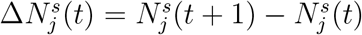, and Δ*P*_*j*_(*t*) = *P*_*j*_(*t* + 1) − *P*_*j*_(*t*). The meaning of each parameter is given in Table 1. Also, we assume that the recovered and confirmed individuals would stay in region *j*.

**Table 1:** Parameters of the Migration-Data Augmented SEICR model.

|  |  |
| --- | --- |
| $\alpha_j$: | rate of infecting a susceptible individual by an exposed individual in region $j$ |
| $\beta_j$: | rate of infecting a susceptible individual by an infected individual in region $j$ |
| $\rho_j$: | rate of infecting a susceptible individual by a confirmed individual in region $j$ |
| $\lambda_j$: | confirmed rate of infected individuals in region $j$ |
| $\kappa$: | rate of an exposed individual becoming infected |
| $\gamma_j^{(I)}$: | recovery rate of an infected but not confirmed individual in region $j$ |
| $\gamma_j^{(Q)}$: | recovery rate of a confirmed individual in region $j$ |
| $k_I$: | possibility of an infected individual moving from one region to another |
| $k_j^{(c)}$: | increase in number of individuals eventually infected for each additional infected or exposed individual in region $j$ |
| $\psi_j$: | level of active intervention, $\psi_j = 1/k_j^{(c)}$ |
| $k_h$: | proportion of population infected achieving no further spreading, i.e., absolute upper bound for $N_j^s$ for all $j$ |
| $\delta_j$: | initial percentage of eventual infected individuals in region $j$ |
| $I_{j,0}$: | initial number of infected individuals in region $j$ |
| $E_{j,0}$: | initial number of individuals in region $j$ |
| $C_{j,0}$: | initial number of confirmed infected individuals in region $j$ |

In this SEICR model, the number of individuals eventually infected is set initially at 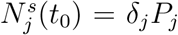(*δ*_*j*_ being constant), implying that some effective measures have been taken by the authorities to limit the upper bound of the susceptible population. Moreover, in the case of inactive or less effective intervention, or even unchecked spread, the growth of the number of infected cases will add to the eventual infected number. Hence, the number of eventually infected individuals should increase for each additional infected or exposed individual at time *t*. This is equivalent to adding an extra term (the boxed term below) to Δ*S*_*i*_(*t*) and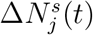. Furthermore, as the number of infected cases increases and approaches a saturating percentage *k*_*h*_ (such as a herd-immunity condition), the spreading is expected to slow down significantly, i.e., *α*_*j*_ and *β*_*j*_ will drop as *N*_*s*_ approaches *k*_*h*_*P*_*j*_, where 0 *< k*_*h*_ *<* 1. Thus, we have

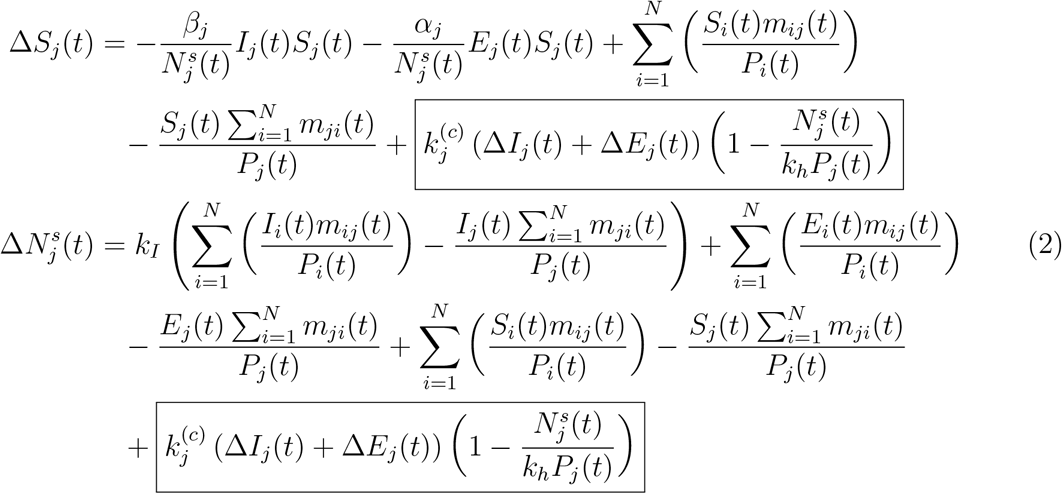

where 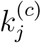 is an inverse indicator of the level of active intervention implemented, and corresponds quantitatively to an increase in the number of eventual infected individuals for each additional infected or exposed individual in region *j*, and the added term in Δ*S*_*j*_ and 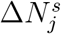 will approach zero as 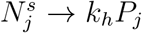. The meanings of other parameters are given in Table 1. Again, the recovered and confirmed individuals are assumed to stay in region *j*.

The model given in (1) and (2) is very general in the sense that it applies to populations with varied levels of effectiveness of active intervention during the outbreak. To further facilitate the assessment of control measures implemented in region *j*, we define the level of active intervention as

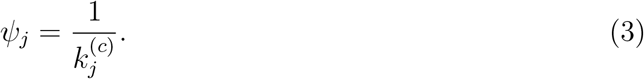

Thus, if *ψ*_*j*_ *>* 1, the control measures are effective and the progression is limited such that 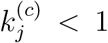. The total number of eventually infected individuals is equal to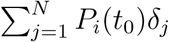.

However, in the case of less effective or ineffective control, i.e., *ψ*_*j*_ *<* 1, infected and exposed individuals continue to spread the disease, and for each additional infected individual, there will be 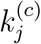 more eventual infected individuals, and the pandemic progresses until the number of infected cases reaches *k*_*h*_*P*_*j*_.

### 3.2 Parameter Identification

The model represented by (1) and (2) describes the dynamics of the pandemic propagation with consideration of human migration dynamics and the reality of insufficient testing that leads to confirmed infected cases being fewer than the actual infected cases. The parameters in (1) and (2) are unknown and to be estimated from historical data of *C* and *R*. We solve this parameter identification problem via constrained nonlinear programming (CNLP), with the objective of finding an estimated growth trajectory that fits the data. An estimated number of infected cases of each city can be generated from (1) and (2) with unknown set *θ*_*j*_ given by

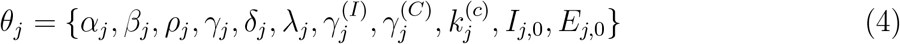

where *I*_*j*,0_ = *I*_*j*_(*t*_0_) and *E*_*j*,0_ = *E*_*j*_(*t*_0_) are the initial numbers of infected and exposed individuals in region *j*, and 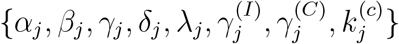 are the model parameters of region *j*. Here, we assume that all confirmed cases are either quarantined or hospitalized, and hence not infectious, i.e., *ρ*_*j*_ = 0. Then, the unknown set is Θ ={*θ*_1_, *θ*_2_,, *θ*_*K*_, *κ, k*_*I*_, *k*_*h*_}, which essentially has 8*K*+3 unknowns, where *K* is the number of regions in the entire population under study. The identification of unknown parameters would require a considerable effort of computation.

Specifically, the parameter estimation problem can be formulated as the following constrained nonlinear optimization problem:

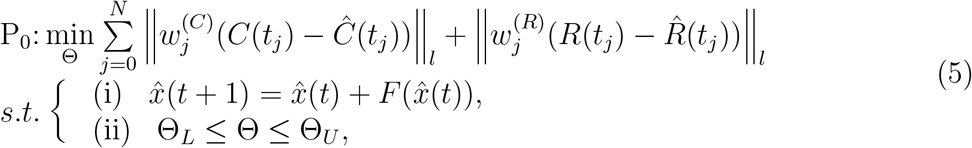

where *F* (*·*) represents the model given by (1) and (2), 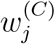 and 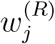 are the weighting coef-ficients, and 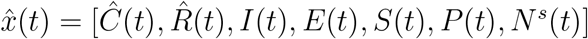 is the set of estimated variables, with unknown set Θ being bounded between Θ_*L*_ and Θ_*U*_. In this work, an inverse approach is taken to find the unknown parameters and states by solving (5).

### 3.3 Prediction

The model parameters characterize the spreading dynamics, and once the set of parameters has been identified using the abovementioned optimization procedure, we may generate future progression profiles by using the same set of parameters. Moreover, we may also adjust some of the parameters to examine different possible scenarios, corresponding to varying levels of active intervention 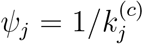, including travel restriction, mandatory quarantine and other control measures. If the level of active intervention stays with the status quo, we will use the same value of 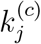 for generating future progression profiles. Future paths under more active intervention can be predicted by reducing the value of 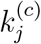. In our study, we will consider three levels of active intervention, namely,

1. staying with the status quo, corresponding to the same value of *ψ*_*j*_ or 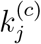;
2. two-fold step-up of active intervention, corresponding to 2*ψ*_*j*_ or 0.5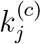;
3. four-fold step-up of active intervention, corresponding to 4*ψ*_*j*_ or 0.25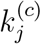.

We will examine the results in the next section.

## 4. Results

### 4.1 Parameters and Prediction for Japan

The pandemic progression profiles of 47 Japanese prefectures or regions are examined. We perform data fitting of the model, described by (1) and (2), using historical daily data of confirmed and recovered cases [11, 12]. A typical candidate set of parameter values that fit well wth the data from January 15, 2020 to March 20, 2020 is as follows:

- 1.3941 *<* 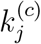*<* 1.5979; 0.0982 *< α*_*j*,0_ *<* 0.1158; 0.3895 *< β*_*j*,0_ *<* 0.5163; 0.0098 *<* 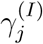 *<* 0.0128; 0.0027 *<*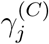 *<* 0.0047; 0.0019 *< λ*_*j*_ *∈<* 0.0052; *κ* = 0.1861; *k*_*h*_ = 0.6514.

This set of parameters reflects an inadequate level of control to slow the spread of the disease, as indicated by the value of 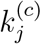 being larger than 1. Specifically, for each additional infected or exposed individual, the number of eventual infected individuals would increase by around 1.5 on the average. The number of individuals eventually infected will approach a saturating percentage *k*_*h*_.

We have identified 100 candidate sets of parameters which satisfy the fitting criteria, and for each set of parameters, we perform a separate simulation run. Figure 2 shows one particular simulation run of a well fitted candidate set of parameters for 8 selected prefectures in Japan. The averaged results of all simulation runs are consolidated in the charts shown in Figure 2. Based on the data up to March 20, 2020, our model estimates that less than 3% of the infected cases are confirmed, with Hokkaido having the highest percentage (6.9%) and Hyogo-ken the least (1.5%), as shown in Figure 3(a). In other words, the actual number of infected individuals could be 20 times as many as the official confirmed number. Statistics of percentages of the population confirmed and infected with the disease up to March 20, 2020 are shown in Figure 3(b).

**Figure 2:**
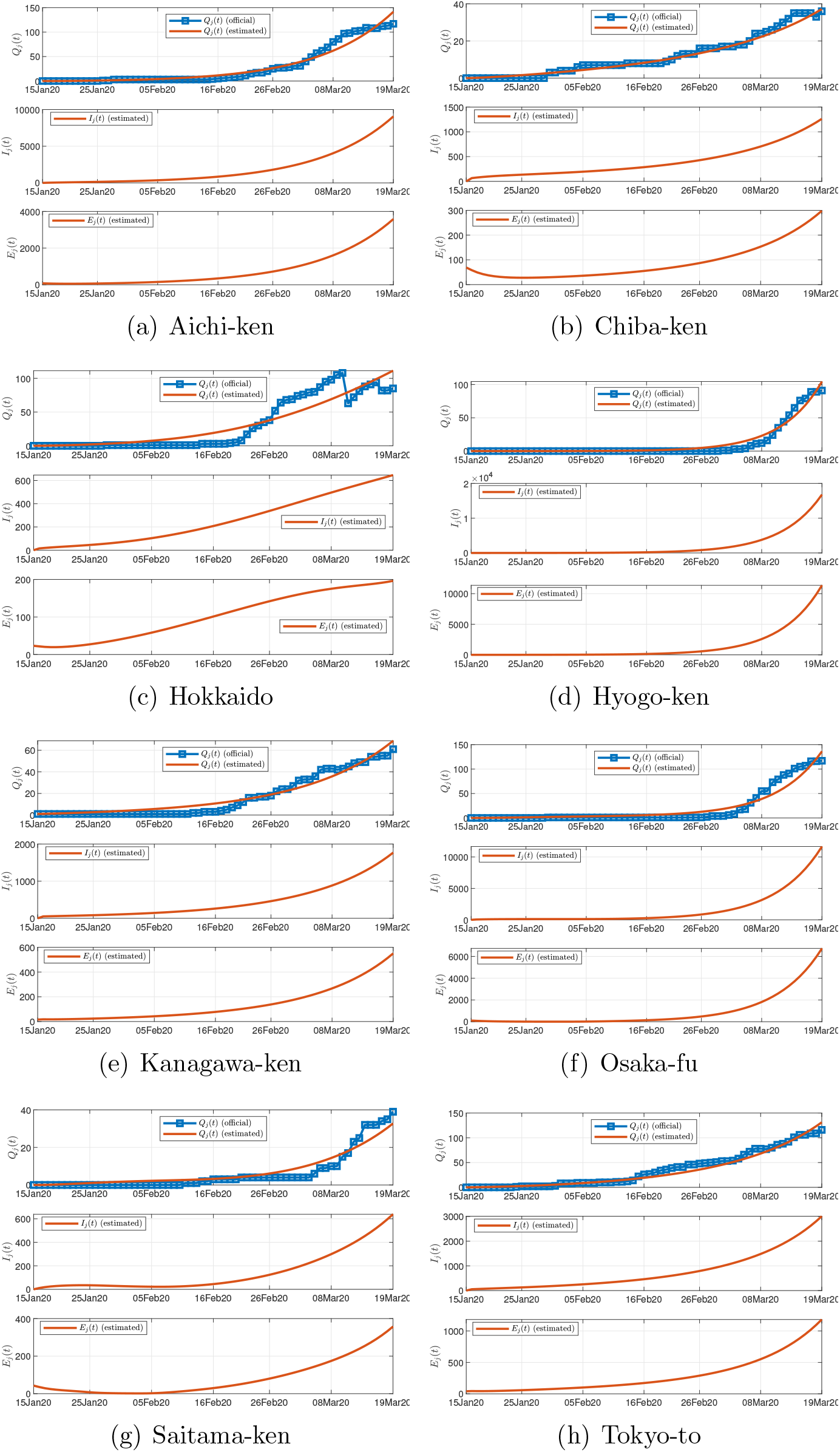
Official and estimated number of confirmed infected individuals in 8 selected prefectures in Japan (upper), estimated number of infected individuals (not confirmed) (middle), and estimated number of exposed individuals (lower).

**Figure 3:**
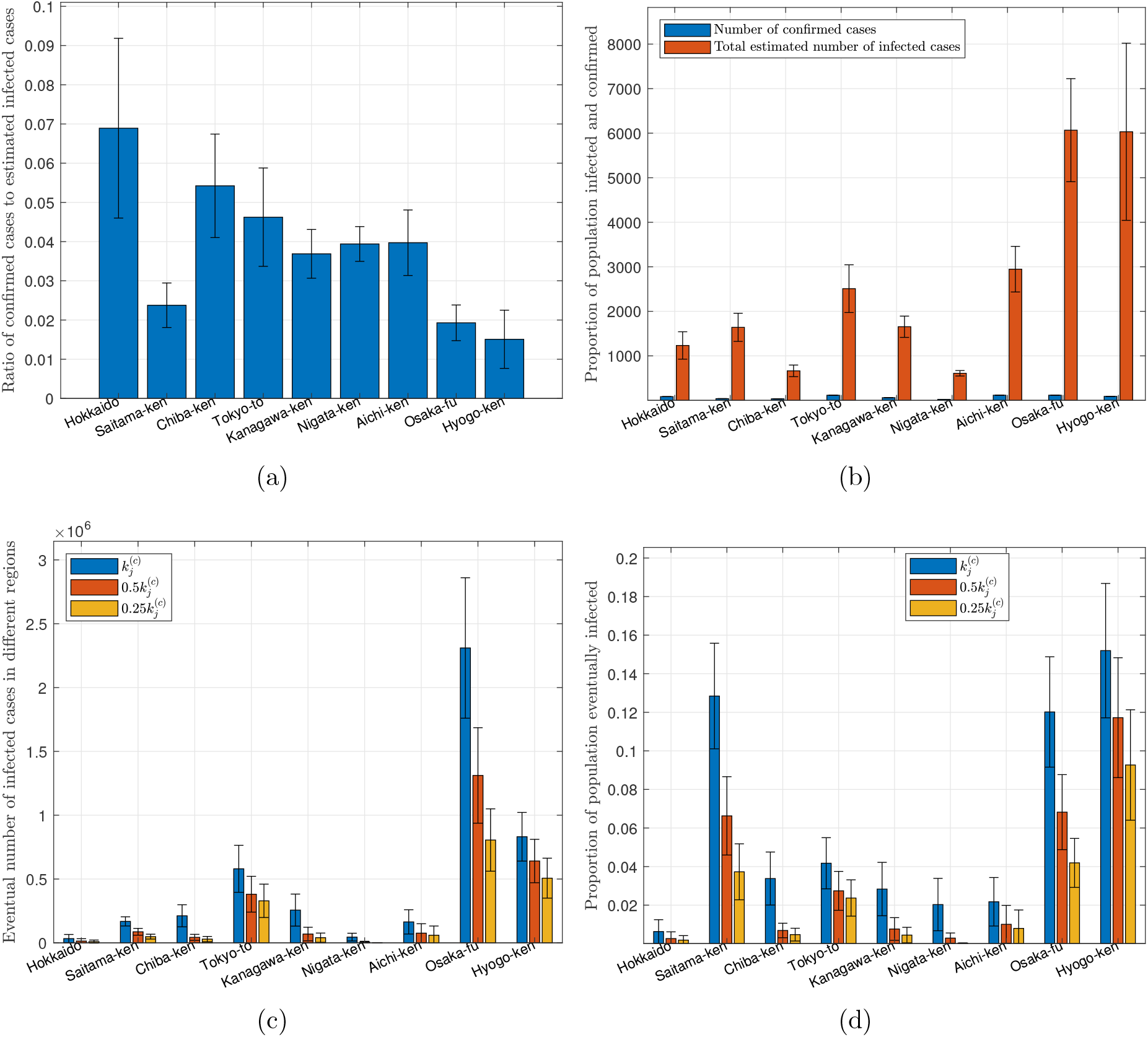
Statistics of data and predicted results for Japan. (a) Proportion of infected cases that are confirmed as of March 20, 2020; (b) number of confirmed cases and estimated number of infected cases as of March 20, 2020; (c) number of individuals eventually infected under three levels of active intervention;(d) proportion of population eventually infected under three levels of active intervention.

By extending each simulation run to the forthcoming 200 days, we obtain a set of predicted progression profiles for each region in Japan. Moreover, different levels of active intervention can be assessed by adjusting parameter 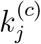 relative to the values found in each candidate set. For instance, by reducing 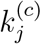 and re-running the simulation, we may assess the effect of tightening the control measures. Specifically, as described in Section 3.3, we examine three cases, corresponding to the level of active intervention being unchanged, two-fold elevated and four-fold elevated. The key results are summarized as follows:

- Staying with the status quo (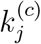 unchanged): If there is no further tightening of control aiming to slow the spread, all parameters of the candidate sets will remain unchanged. The total number of individuals eventually infected until September 23, 2020 in each region is shown in Figure 3(c). In this case, the number of infected individuals in Osaka-fu and Tokyo-to will reach about 2,300,000 and 600,000 (12% and 4.2% of population), respectively, while most other regions will have around 5% of the population eventually infected by September 23, 2020, as shown in Figure 3(d). In total, about 6.55% of the population in Japan will be infected.
- Two-fold elevated active intervention (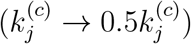: If active intervention is stepped up to twice the current level, i.e., the value of 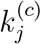 set to half of the original value in each simulation run, we observe a significant drop in the number of individuals eventually infected, as given in Figure 3(c). Specifically, the percentage of population eventually infected by September 23, 2020 in Osaka-fu and Tokyo-to would drop to about 6.8% and 2.3%, respectively, while most other regions would drop to less than 2%, as shown in Figure 3(d). In total, about 4.14% of the population in Japan will be infected.
- Four-fold elevated active intervention (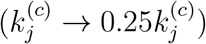: If active intervention is stepped up to four times the current level, i.e., the value of 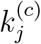 set to a quarter of the orig-inal value in each simulation run, we observe a very drastic drop in the number of individuals eventually infected, as given in Figure 3(c). Specifically, the percentage of population eventually infected by September 23, 2020 in Osaka-fu and Tokyo-to would drop to about 4.1% and 2.3%, respectively, while most other regions would drop to less than 1%, as shown in Figure 3(d). In total, about 1.54% of the population in Japan will be infected.

In conclusion, the current level of control by the Japanese government seems to be inadequate, and a significant step-up in the level of active intervention is necessary in order to curb the aggressive progression trend. In addition, our model estimates that the number of infected individuals could be 20 times as many as the currently confirmed number due to various reasons such as insufficient testing. Based on the data collected so far and assuming no further tightening of control, our model estimates about 6.65% of the population eventually infected, and a four-fold elevation in control efforts may bring it down to 1.54% (about 75% reduction) and end the pandemic sooner. As will be shown in Section 4.3, a drastic 10-fold elevated active control may bring it further down to 0.24%.

### 4.2 Parameters and Prediction for the USA

The pandemic progression profiles of 50 states and a federal district in the USA are examined. We again perform data fitting of the model, described by (1) and (2), using historical daily data of confirmed and recovered cases from February 20 to March 20, 2020 [11, 12], and obtain 100 candidate sets of parameters that satisfy the fitting criteria. For brevity of presentation, we show here the results for eight selected states having significant numbers of infected individuals as of March 20, 2020. Figure 4 shows one typical simulation run, showing the number of confirmed cases, the estimated number of infected individuals (not confirmed), and the estimated number of exposed individuals.

**Figure 4:**
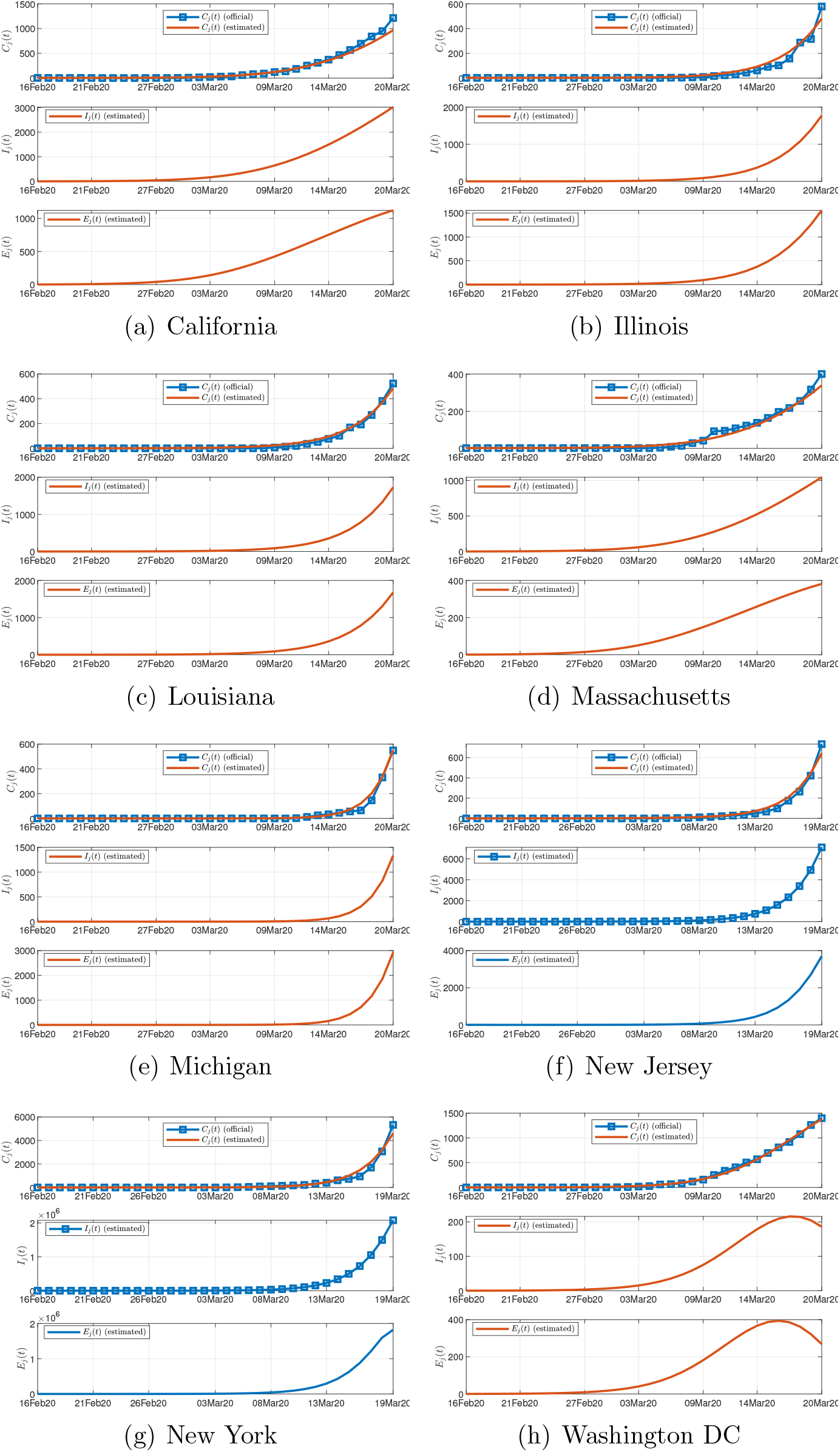
Official and estimated number of confirmed infected individuals in 8 selected states in USA (upper), estimated number of infected individuals (not confirmed) (middle), and estimated number of exposed individuals (lower).

As of March 19, 2020, our model shows that less than 20% of the infected cases are confirmed, with Washington DC having the highest percentage (36%) and Michigan state the least (0.7%), as shown in Figure 5(a). In other words, the actual number of infected individuals in the USA could be 5 times as many as the confirmed number. Statistics of percentages of population confirmed and infected with the disease up to March 19, 2020 are shown in Figure 5(b).

**Figure 5:**
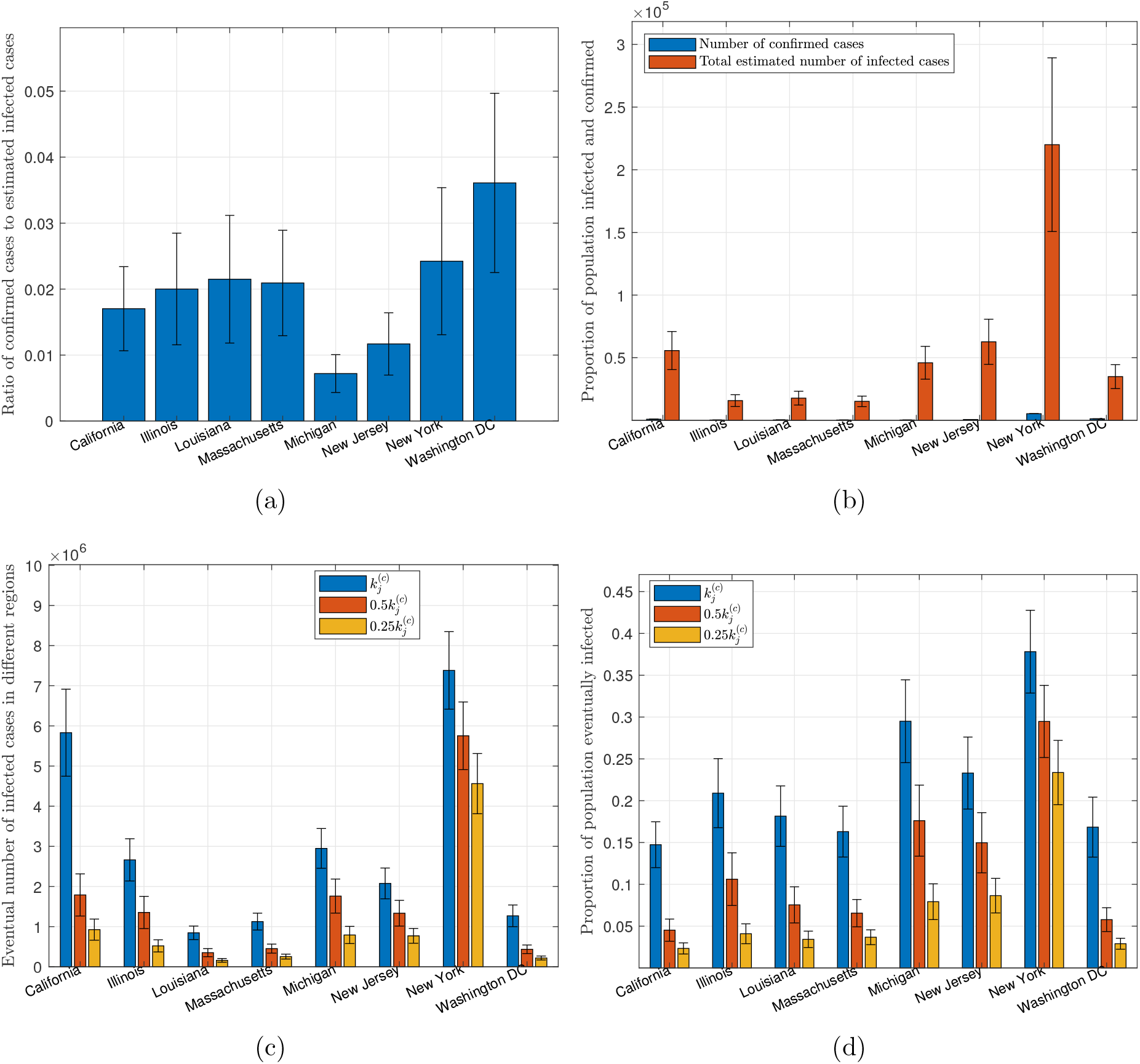
Statistics of data and predicted results for USA. (a) Proportion of infected cases that are confirmed as of March 19, 2020; (b) number of confirmed cases and estimated number of infected cases as of March 19, 2020; (c) number of individuals eventually infected under three levels of active intervention; (d) proportion of population eventually infected under three levels of active intervention.

Again, by extending each simulation run to the future 200 days, we obtain a set of predicted progression profiles for each state and federal district in the USA. We also examine three cases corresponding to three different levels of active intervention, as described in Section 3.3. The key results are summarized as follows:

- Staying with the status quo (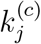 unchanged): If there is no further tightening of control aiming to slow the spread, all parameters of the candidate sets will remain unchanged. The total number of individuals eventually infected until September 23, 2020 in each state is shown in Figure 5(c). In this case, the number of infected individuals in California and New York state will reach about 5,800,000 and 7,300,000 (15% and 37.5% of population), respectively, while most other states will have less than 20% of the population eventually infected by September 23, 2020, as shown in Figure 5(d). In total, about 18.2% of the population in the USA will be infected.
- Two-fold elevated active intervention (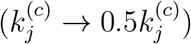: If active intervention is stepped up to twice the current level, i.e., the value of 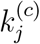 set to half of the original value in each simulation run, we observe a significant drop in the number of individuals eventually infected, as given in Figure 5(c). Specifically, the percentage of population eventually infected by September 23, 2020 in California and New York state would drop to about 4.5% and 29.5%, respectively, while most other states would drop to less than 10%, as shown in Figure 5(d). In total, about 14% of the population in the USA will be infected.
- Four-fold elevated active intervention (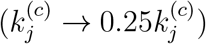: If active intervention is stepped up to four times the current level, i.e., the value of 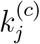 set to a quarter of the original value in each simulation run, we observe further reduction in the number of individuals eventually infected, as given in Figure 5(c). Specifically, the percentage of population eventually infected by September 23, 2020 in California and New York state would drop to about 2.5% and 23%, respectively, while most other states would drop to less than 3%, as shown in Figure 5(d). In total, about 9.32% of the population in the USA will be infected.

In summary, a significant step-up in the level of active intervention is necessary for US government to slow the spread of the virus. In addition, our model estimates that the number of infected individuals could be five times as many as the currently confirmed number due to insufficient testing and other reasons. Based on the data collected so far and assuming no further tightening of government’s control, our model estimates that about 18.2% of population would eventually be infected, and a four-fold elevation in control efforts may bring it down to 9.32%. As will be shown in Section 4.3, a drastic 10-fold elevated active control may bring it further down to 5.24%.

### 4.3 Further Control

The results presented above have highlighted the ability of the model in assessing the impact of active intervention through adjusting one of the parameters, namely, 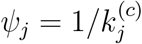. Moreover, it has been widely disseminated that maintaining personal hygiene is equally important in curbing the spread of the virus. The World Health Organization recommends several specific protective measures to be practiced by the public, including frequent hand washing, maintaining social distancing, avoiding touching one’s eyes, nose and mouth, and practicing respiratory hygiene [16]. Recent studies also show that wearing surgical masks also help in some cases [17, 18]. The level of vigilance of the public in exercising personal protective measures can also be incorporated in our model through adjusting infection rates *α*_*j*_ and *β*_*j*_. We can therefore assess the combined effectiveness of active intervention and practicing protective measures in controling the pandemic. Here, we vary *α*_*j*_, *β*_*j*_ and 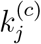 from 10% to 100% of the originally identified values in 10 intervals, corresponding 10 different levels of vigilance of the public and active intervention by the authorities. In particular, we assess *α*_*j*_ and *β*_*j*_ as one property and 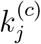 as another, i.e., varying *α*_*j*_ and *β*_*j*_ in synchrony. Specifically, for each candidate parameter set, we perform 100 simulation runs for each combination of *α*_*j*_, *β*_*j*_ and 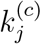, where *α*_*j*_, *β*_*j*_ and 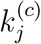 vary from 10% to 100% of the original values in 10 steps. Then, we investigate the percentage of population eventually infected in Japan and the USA. Results are shown in Figures 6(a) and 6(b).

**Figure 6:**
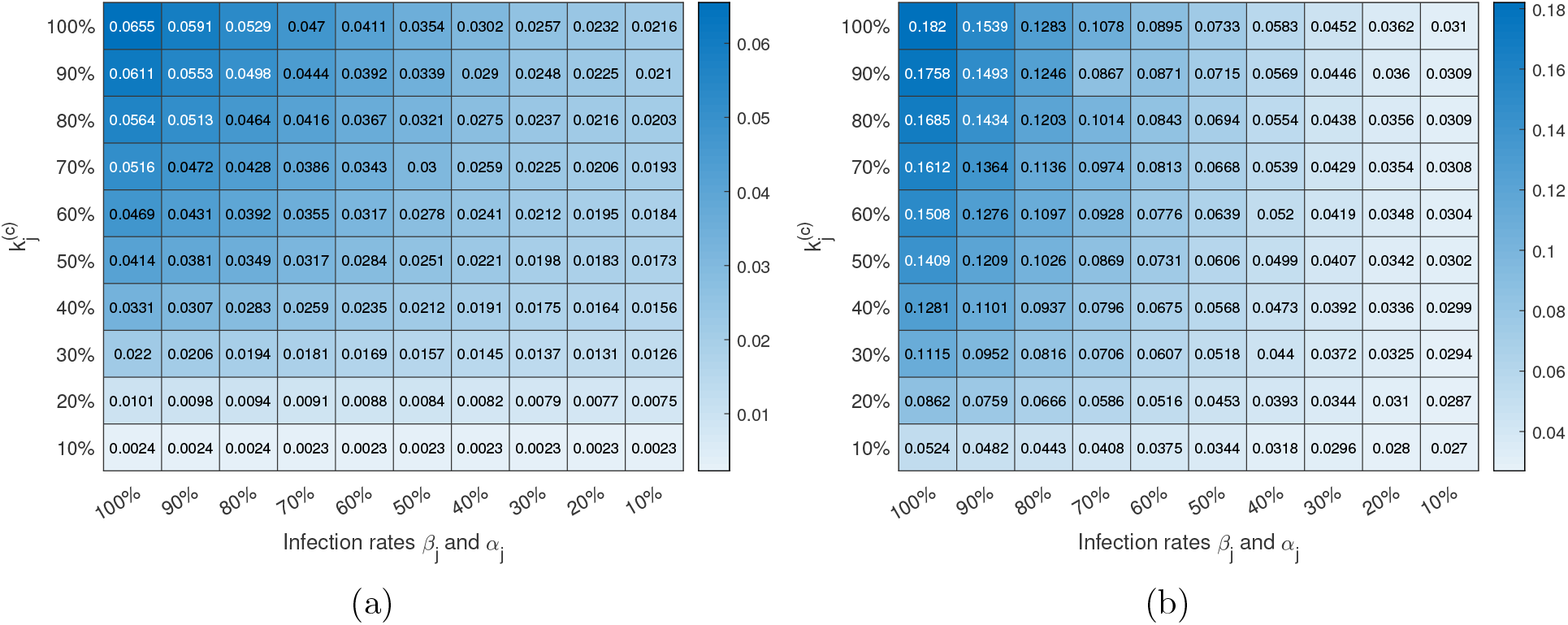
Proportion of population eventually infected under different levels of active intervention indicated by 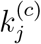 (smaller the stronger) and maintaining personal hygiene and exercising protective measures indicated by *α*_*j*_, *β*_*j*_ (smaller the stronger). (a) Japan; (b) USA.

The mean percentage of population eventually infected under different combinations of parameter values for Japan and the USA are given in the charts shown in Figures 6(a) and 6(b), respectively. For instance, suppose the level of vigilance of the public has dramatically raised and the level of active intervention has been stepped up, resulting in 90% reduction in the infected rates and 90% reduction in 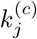, i.e., parameters changed to 0.1*β*_*j*_, 0.1*α*_*j*_, and 0.1 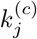. Referring to Figures 6(a) and 6(b), the percentage of population eventually infected can be dramactcially educed to 0.23% for Japan and 2.7% for the USA. Similar interpretations can be taken for any other combination of levels of public vigilance and active intervention.

Our results have highlighted an interesting difference between the effectiveness of government’s active intervention and maintaining personal hygiene by the public for Japan and the USA. For Japan, we observe a 27-fold reduction (from 6.55% to 0.24%) in the percentage of individuals eventually infected upon a drastic 10-fold step-up of active intervention, as given in the first column of Figure 6(a), whereas less than 3-fold reduction (from 6.55% to 2.16%) is observed in the percentage of individuals eventually infected upon the same 10-fold improvement in personal hygiene, as shown in the first row of Figure 6(a). Thus, government’s active intervention seems to be more important for Japan. Moreover, for the USA, we see the opposite. Specifically, only about 4-fold reduction in the percentage of individuals eventually infected is observed upon a drastic 10-fold step-up of active intervention, as shown in the first column of Figure 6(b), whereas a 6-fold reduction is observed upon a 10-fold improvement in maintaining personal hygiene by the public. Thus, raising the level of vigilance of the public in exercising personal protective measures is comparatively more important for the USA. A plausible reason for the difference between Japan and USA is that the model has captured higher infection rates for the USA compared to Japan. Reducing *k*_*j*_ for the US case is thus less effective at such high infection rates, i.e., a fewer eventual infected number per additional infected individual would not help too much. In contrary, the parameter sets for Japan already have relatively lower infection rates, and further improvement by reducing the infection rates would be limited.

As a final remark, we observe from the charts given in Figure 6 that combining a very high level of vigilance of the public in exercising strict protective measures and a drastic step-up of government intervention, the percentage of the population getting infected can be reduced to 0.23% in Japan and 2.7% in the USA.

## Conclusion

One of the key challenges in data-driven modeling and analysis is the delayed and missing information that makes fitting of models either difficult or unreliable, resulting in inconsistent or even erroneous dynamical profiles generated by a poorly parameterized model. The traditional Susceptible-Exposed-Infectious-Recovered (SEIR) model provides a general dy-namical description of the spreading of a disease in a population, and involves a series of transitional processes that describe how a healthy individual becomes exposed, infected and eventually recovered or removed from the population. The model thus generally has four dynamical states. From the modeling point of view, the model parameters can be extracted from fitting with historical data consisting of the number of individuals in the infected state and recovered state, which are the usual practically observable data. Outbreaks of the 2019 New Coronavirus (COVID-19) pandemic have occurred in over 185 countries since the virus began to spread from China in January 2020 via an active global transportation network. The data of infected and recovered cases reported by different cities and regions have been found unreliable or incomplete, as they are subject to the availability of test facilities as well as other factors related to bureaucracy of reporting and the mode of operation of the medical systems. Nonetheless, figures of “confirmed” cases can still be honest figures, though not necessarily the true figures of infected cases.

In this work, we propose a new disease spreading model with consideration of the delayed and missing data of infected cases, intercity travel, and the level of active intervention. Instead of matching the number of confirmed cases obtained from official sources directly with the number of infected cases in the model, we create a new state which is a delayed and contracted version of the original infected state of the SEICR model, leading to a new SEI**C**R model. The model, which estimates the actual number of infected cases after identifying the best parameter sets, is applied to study the COVID-19 pandemic progression in Japan and the USA. Results reveal that the actual number of infected individuals could be up to 20-fold and 10-fold as many as the confirmed numbers in Japan and the USA, respectively, as of March 19, 2020. Our model also allows assessment of varying levels of active intervention implemented by the government, and results show that the current level of control by the Japanese and US governments may be inadequate, and a significant step-up in the level of active intervention is necessary in order to slow the aggressive progression trend in both countries. For Japan, based on the data collected so far and assuming no further tightening of control, our model estimates about 6.55% of the population eventually infected, and a four-fold elevation in control efforts may bring it down to 1.54%. For the USA, our model estimates about 18.2% of population eventually infected if the government does not step up its control, and a four-fold elevation in active intervention may bring it down to 9.32%. Finally, adjusting the infection rates permits assessment of effectiveness of practicing protective measures and maintaining personal hygiene. Simulations of various levels of implementation of combined active intervention and protective measures show that stepping up government’s active intervention would be more effective for Japan, while raising the level of vigilance of the public in maintaining personal hygiene and social distancing is comparatively more important for the USA. With the public raising its level of vigilance and the government drastically elevating its active intervention, the percentage of population getting infected can be reduced to 0.23% in Japan and 2.7% in the USA.

## Data Availability

All data used in this paper are available at the links given below.

https://en.wikipedia.org/wiki/2020\_coronavirus\_pandemic\_in\_Japan

https://www.who.int/emergencies/diseases/novel-coronavirus-2019

https://www.e-stat.go.jp/dbview?sid=0003280293

https://www.transtats.bts.gov

## Acknowledgment

This work was supported by National Science Foundation of China Project 61703355, Guangdong Youth University Innovative Talents Project 2016KQNCX223, and City University of Hong Kong under Special Fund 9380114.

